# Internet Addiction and its Associated Factors Among Undergraduate Students of Institute of Medicine, Nepal

**DOI:** 10.64898/2026.09.07.26362437

**Authors:** Rima Lawaju, Sushan Man Shrestha

**Affiliations:** Central Department of Public Health, Institute of Medicine, Kathmandu, Nepal

## Abstract

Internet use has increased rapidly around the world, along with reports of overuse among internet users. This can affect not just the psychosocial well-being but also the physical health of the young population, who are its major victims. However, very limited studies have been conducted on internet addiction among the young population in Nepal. This study aims to find the prevalence of internet addiction and its associated factors among undergraduate students of the Institute of Medicine (IOM), Nepal. We conducted a cross-sectional study among 211 undergraduate students of IOM. A structured questionnaire was used to collect socio-demographic, academic, and internet-use-related information. Internet addiction was assessed using Young’s Internet Addiction Test. Descriptive and bivariate analyses were performed. Frequencies and percentages were calculated for univariate analysis, while bivariate analysis was performed to assess the association between the dependent and independent variables. One in two students had internet addiction, with 36.5% having mild addiction, 12.3% having moderate addiction, and 1.4% having severe addiction. The mean age (±SD) of the students was 21.6 ± 2.0 years. The variable found to be associated with internet addiction was the duration of internet use per day. Students who used the internet for more than five hours per day were about 2.3 times more likely to have internet addiction than those who used the internet for five hours or less (OR = 2.399, 95% CI: 1.334–4.314). Internet addiction is a significant concern among undergraduate students and may increase if not appropriately addressed. Excessive and problematic internet use may contribute to mental health problems such as difficulty concentrating, anxiety, and compulsive behaviors, particularly among young people who are frequent users of the internet.

## Introduction

The most widely used method of exchanging information, communicating, having fun, and conducting business is the internet. Internet has countless benefits and advantages in our new technological era. However, there is more and more research and written works about the drawbacks of its excessive and pathological use (1). The Internet has developed into a limitless arena for information sharing, social networking, and the growth of cyber habits influencing our mental and social welfare. It is no longer merely an infrastructure (2).

Internet addiction (IA), according to Chou et al., is the use of the Internet in a way that causes psychological, social, academic, and professional problems (3). Addiction to problematic internet use is characterized by excessive or unchecked cravings, preoccupations, or behaviors related to Internet use that cause impairment or suffering. Addicts may spend a lot of time online, cut themselves off from other social interactions, and pay little attention to bigger-picture events in their lives. They spend numerous hours each week on the Internet, primarily employ dysfunctional coping mechanisms, and exhibit weaker interpersonal relationships than peers who do not display any indicators of problematic internet use (4). The idea of IA, which was introduced by Goldberg in 1995, has recently become well-known. Various labels, including “net addiction,” “online addiction,” “IA disorder,” “pathologic internet use,” and “cyber disorder,” are used to describe this problem. Young modified the DSM-IV criteria to correspond to internet use in the Internet Addiction Test (IAT) and most closely correlated excessive internet use with pathological gambling, an impulsivity disorder (5).

A cross-sectional, cross-cultural study conducted among adults across nine European countries reported prevalence rates of 14–55% for moderate to severe internet addiction. Similarly, a population-based epidemiological study conducted among school students across six Asian countries reported prevalence rates of 14–51% for internet addiction (2,6). Two similar studies conducted among university students in Nepal reported that 40% of internet users were mild addicts, 41.53% were moderate addicts, and 3.07% were severe addicts. Similarly, Marahatta et al. reported that 50.8% were mild addicts, 40.7% were moderate addicts, and 1.3% were severe addicts (6,7). Time spent online, patterns of internet use, psychosocial factors, and coexisting symptoms or disorders have been associated with internet addiction. Other factors, including sleep disturbance, lack of physical exercise, and employment status, as well as sociodemographic factors such as age, gender, residence, and relationship status, have also been associated with internet addiction (8).

The number of people accessing the internet globally has increased over the past two decades to approximately 2.5 billion people (7). Nepal has shown extraordinary success in embracing digital technology, with cell penetration nearing 100% and Internet penetration reaching 60% (9). According to one study, Internet addiction is quite common among people ages 18 to 24, who make up the bulk of internet users (10). According to a study, medical students’ high Internet usage is causing them trouble. They don’t get enough sleep, which results in poor mental focus. If this problem is not resolved promptly, it could get worse and reach a higher level of addiction. Globally, internet addiction is becoming a recognized psychological issue. It directly affects stress, anxiety, and depression (11). However, Nepal lacks sufficient research to consider it as a public health threat. Internet addiction, being highly correlated with psychosocial problems, requires early attention and therefore more information regarding its existence in various contexts and population groups. This research attempts to find out the prevalence of internet addiction among undergraduate students of IOM and its associated factors.

## Methods

### Study design and study participants

A web-based descriptive cross-sectional study was conducted in February 2021 among undergraduate students of the Institute of Medicine (IOM), Nepal. The study population included undergraduate students enrolled in various IOM programs, including Bachelor of Medicine and Bachelor of Surgery (MBBS), Bachelor of Dental Surgery (BDS), Bachelor of Ayurvedic Medicine and Surgery (BAMS), Bachelor of Public Health (BPH), Bachelor of Science in Nursing (BSc. Nursing), Bachelor of Pharmacy (B. Pharmacy), Bachelor of Medical Laboratory Technology (BMLT), Bachelor of Optometry (B. Optometry), Bachelor of Science in Medical Imaging Technology (BSc. MIT), Bachelor of Audiology and Speech-Language Pathology (BASLP), and Bachelor in Perfusion Technology.

After obtaining the total number of students and the sampling frame, the students were categorized into six domains as presented in Table 1. The sample size was then calculated proportionately according to the size of each field of study and further stratified by year of study to ensure proper representation of the population.

**Table 1:** Proportionate sample size in different fields of study.

| Field of study | Total no | Proportionate no. |
| --- | --- | --- |
| MBBS/ BDS | 504 | 91 |
| BAMS | 158 | 28 |
| BPH | 158 | 28 |
| BSc Nursing | 156 | 28 |
| B. Pharmacy | 99 | 18 |
| Others (BMLT, BMIT, BASLP, B.Optom and B. Perfusion Technology) | 149 | 27 |
| **TOTAL** | **1224** | **220** |

### Sample size and sampling process

Sample size for the study was calculated using formula; 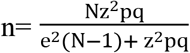, where N=Total number of populations= 1224, Z= level of confidence measure, if α=0.05, Z= 1.96 for 95% CI, p= baseline prevalence of internet addiction. From similar studies, prevalence was taken as 18.88% (6). So, p= 18.88%or 0.1888, q= 1-p= 0.8112, and e=margin of error = 0.05. Using the non-response rate of 10%, the final sample size was calculated as 220. In this study, the final sample size was 211 with a 4.1% non-response rate.

An online survey using a structured questionnaire was conducted. The tool was delivered using Google Forms through email or through social media interaction with the help of class representatives of the respective classes.

### Data collection measures

The instrument of this study was a questionnaire consisting of two parts and 31 questions. The first part collected students’ general information and consisted of 11 items. It included three subcategories: socio-demographic characteristics (age, sex, ethnicity, current residential status, relationship status, and family income), academic characteristics (field of study and year of study), and internet-use-related characteristics (duration of internet use per day, internet speed [time taken to open a webpage during normal use], and purpose of internet use).

The second part of the instrument included Young’s Internet Addiction Test (YIAT), used to assess internet addiction among undergraduates. YIAT consists of 20 items and has a six-point scale, each ranging from 0 to 5. Each statement is weighted along a Likert-scale continuum that ranges from 0 = less extreme behavior to 5 = most extreme behavior for each item. The maximum score is 100 points. A total score of 0 to 30 points is considered to reflect a normal level of Internet usage; scores of 31 to 49 indicate the presence of a mild level of Internet addiction; 50 to 79 reflect the presence of a moderate level; and scores of 80 to 100 indicate a severe dependence upon the Internet (12). Young’s Internet

Addiction Test (YIAT) tool is highly sensitive and highly specific tool with internal consistency of 0.90 (Cronbach"s α) (13).

### Data analysis

Data were entered in Google Forms, and analysis was carried out using IBM SPSS version 21 (IBM, Armonk, NY). Under descriptive statistics, frequencies and percentages were calculated for categorical variables, and means and standard deviations for numerical variables. Further, bivariate analysis was performed to estimate the association between the dependent variable and independent variables.

## Results

The mean age (± standard deviation) of the students was 21.6 ± 2.0 years. Among the students, 61.1% were male. The majority were Brahmin/Chhetri (70.6%), and more than half (56.4%) had been living with their family or relatives for the past six months. Most students were single (82.0%), and 68.7% had a monthly family income above NPR 30,000. Regarding academic characteristics, 39.8% belonged to the MBBS/BDS category, while 33.2% were in fourth year or above. The majority of students (65.9%) used the internet for more than five hours per day, mainly for social networking (32.5%) and entertainment (30.5%). (Table 2)

**Table 2:**
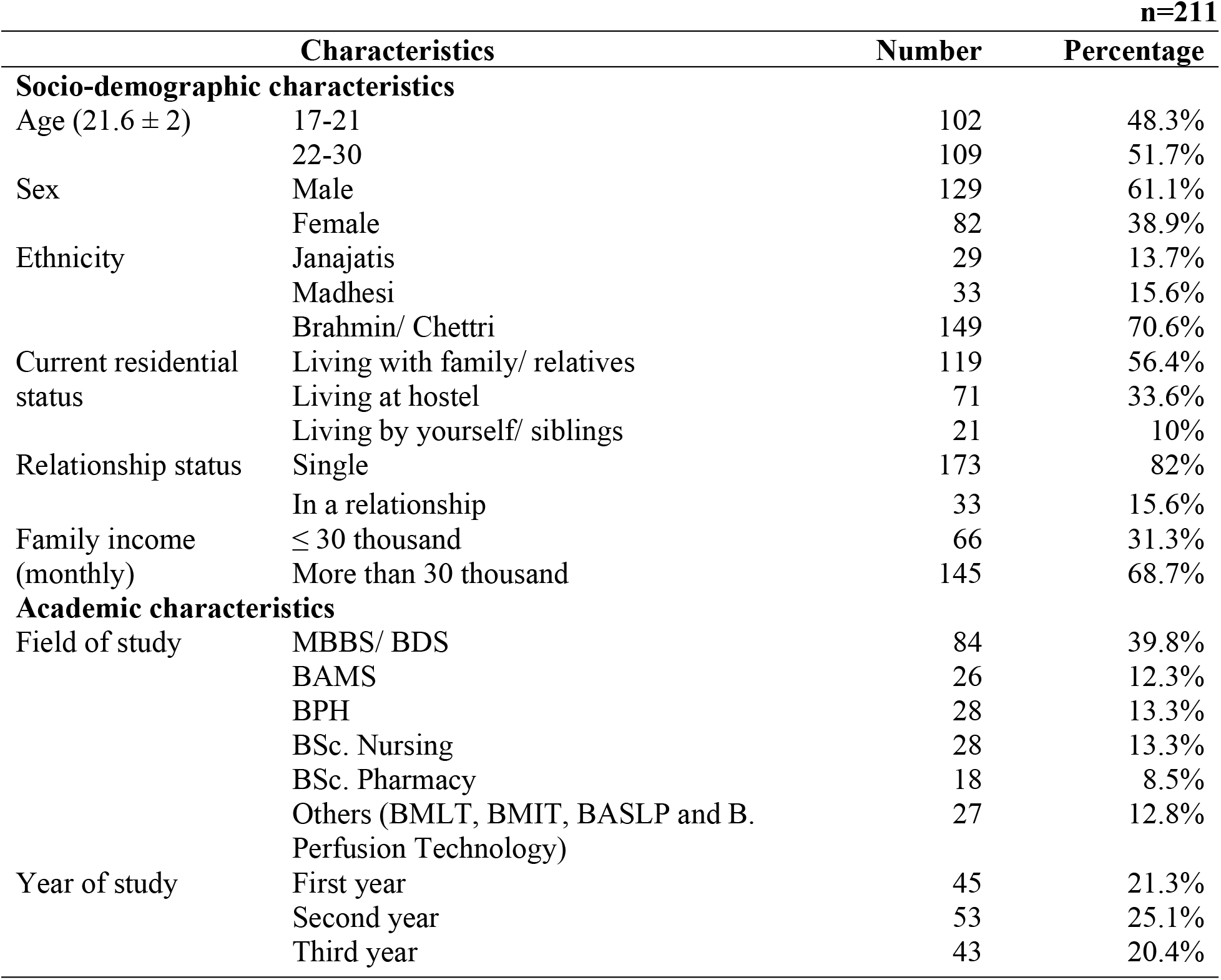
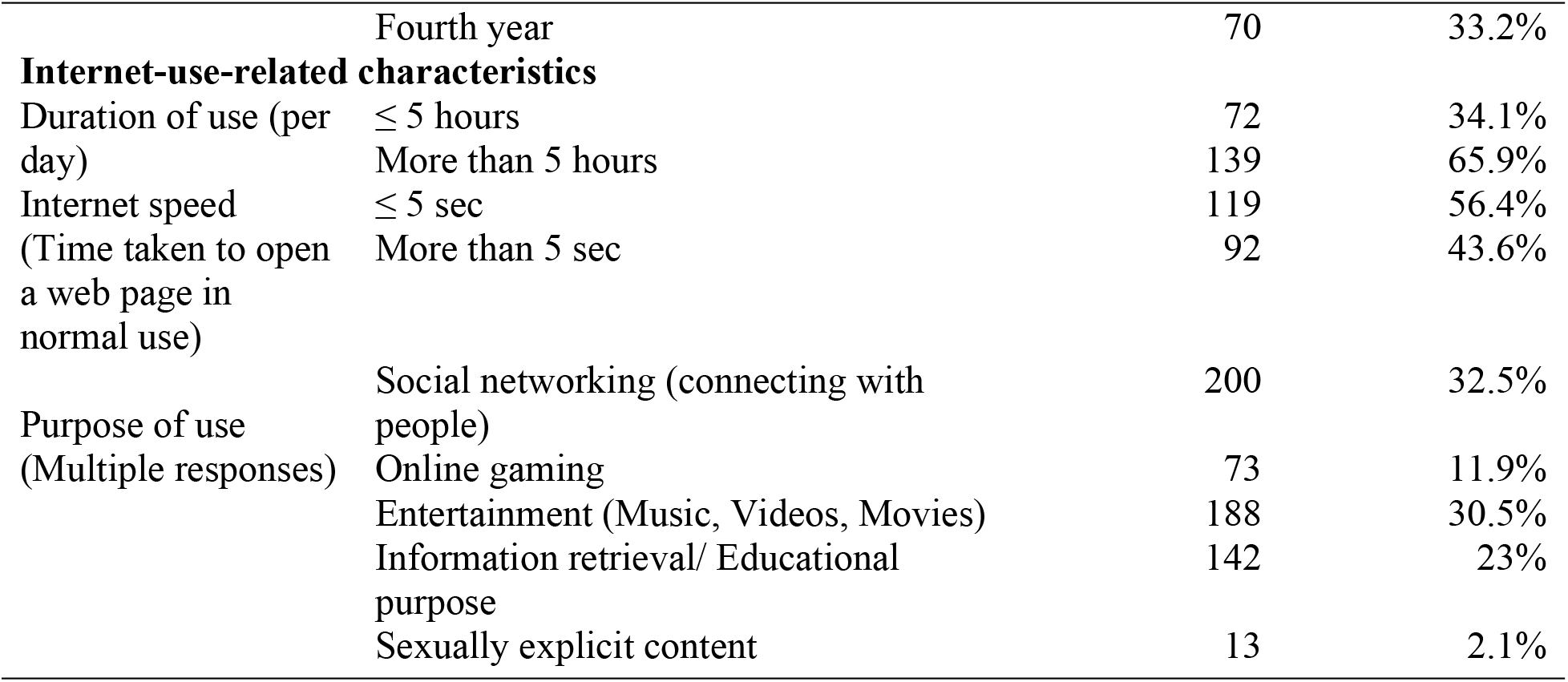
General Characteristics of undergraduate students.

Table 3 presents the internet addiction level of the undergraduate students using Young’s Internet Addiction Test. Nearly half of the students had no internet addiction (49.8%), whereas 36.5% had mild internet addiction, 12.3% had moderate addiction, and 1.4% had severe addiction.

**Table 3:** Internet Addiction level of the undergraduates.

|  |  |  | n=211 |
| --- | --- | --- | --- |
| Level of Addiction | Number | Percentage (%) |  |
| No addiction | 105 | 49.8 |  |
| Mild addiction | 77 | 36.5 |  |
| Moderate addiction | 26 | 12.3 |  |
| Sever addiction | 3 | 1.4 |  |

The bivariate analysis of the association between internet addiction and socio-demographic, academic, and internet-use characteristics of undergraduate students is presented with a 95% confidence interval. None of the socio-demographic variables (age, gender, ethnicity, current residential status, relationship status, and family income) or academic variables (field of study and year of study) were significantly associated with internet addiction. Among the internet-use characteristics, students who used the internet for more than five hours per day were about 2.3 times more likely to have internet addiction than those who used it for five hours or less (OR = 2.399, 95% CI: 1.334–4.314). However, internet speed and purpose of internet use were not significantly associated with internet addiction. (Table 4)

**Table 4:**
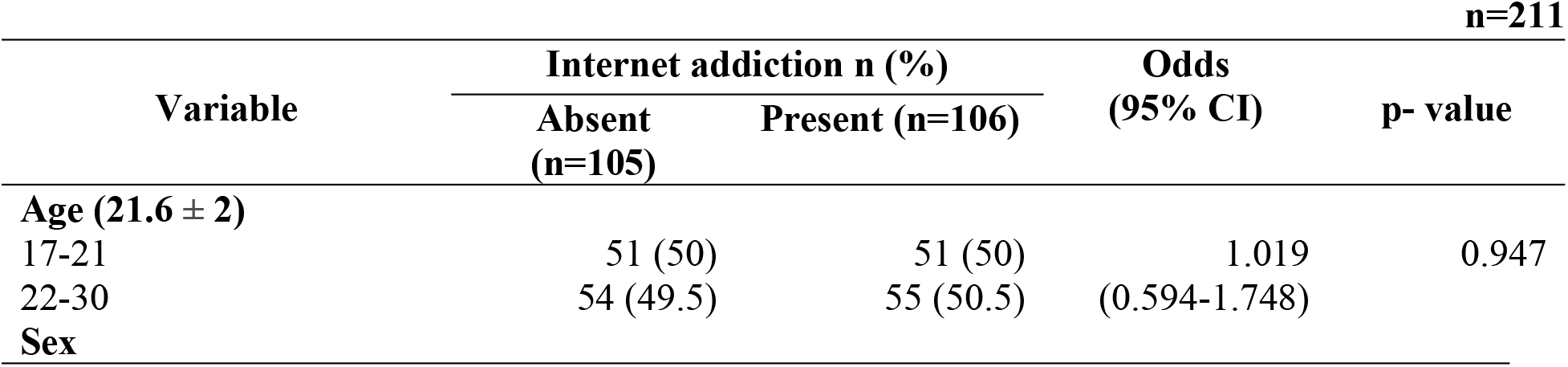
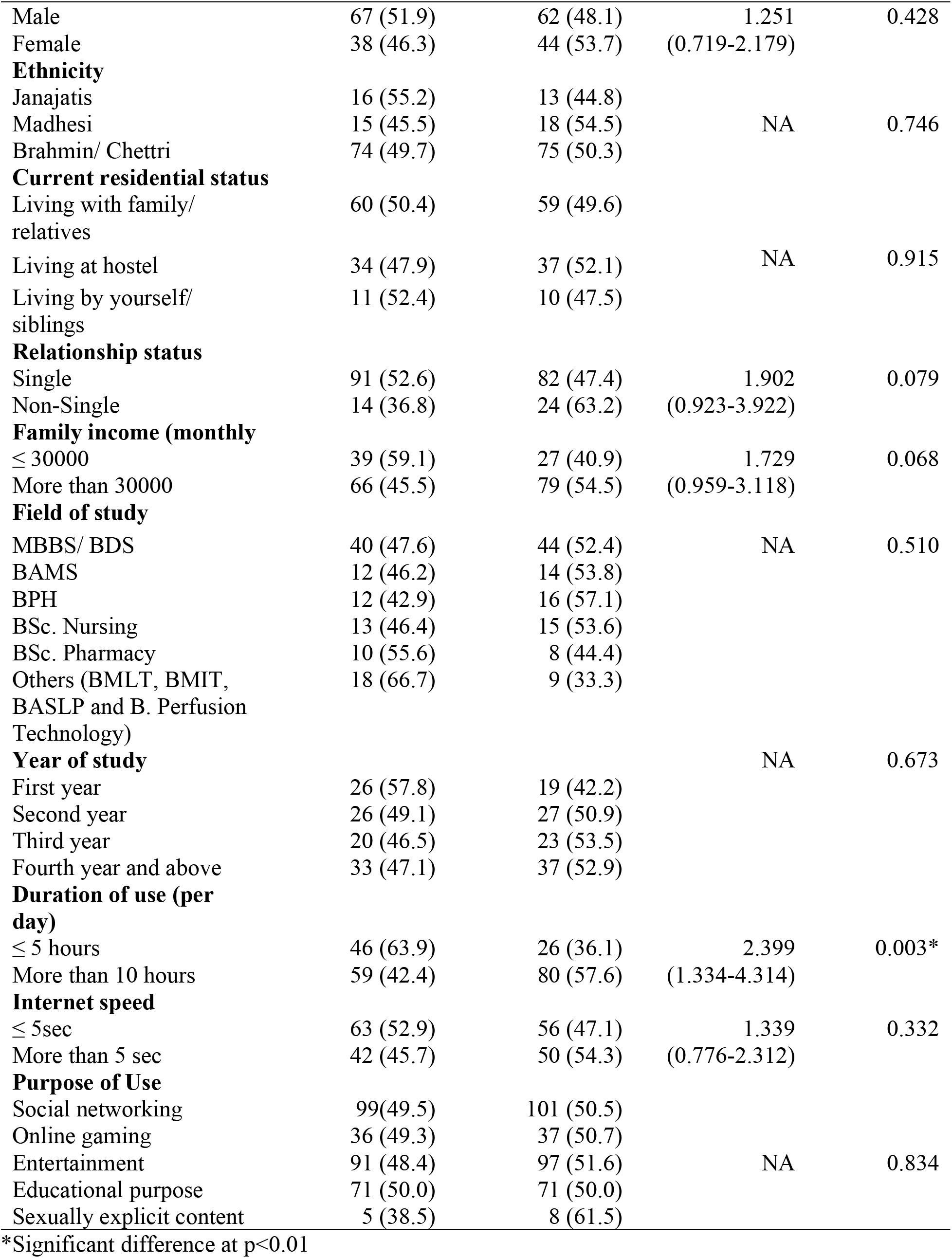
Association between internet addiction and general characteristics.

|  |  |  |  |  | n=211 |
| --- | --- | --- | --- | --- | --- |
| Variable | Internet addiction n (%) |  | Odds (95% CI) | p- value |  |
|  | Absent (n=105) | Present (n=106) |  |  |  |
| **Age (21.6 ± 2)** |  |  |  |  |  |
| 17-21 | 51 (50) | 51 (50) | 1.019 | 0.947 |  |
| 22-30 | 54 (49.5) | 55 (50.5) | (0.594-1.748) |  |  |
| **Sex** |  |  |  |  |  |

|  |  |  |  |  |
| --- | --- | --- | --- | --- |
| Male | 67 (51.9) | 62 (48.1) | 1.251 | 0.428 |
| Female | 38 (46.3) | 44 (53.7) | (0.719-2.179) |  |
| **Ethnicity** |  |  |  |  |
| Janajatis | 16 (55.2) | 13 (44.8) |  |  |
| Madhesi | 15 (45.5) | 18 (54.5) | NA | 0.746 |
| Brahmin/ Chettri | 74 (49.7) | 75 (50.3) |  |  |
| **Current residential status** |  |  |  |  |
| Living with family/<br>relatives | 60 (50.4) | 59 (49.6) |  |  |
| Living at hostel | 34 (47.9) | 37 (52.1) | NA | 0.915 |
| Living by yourself/<br>siblings | 11 (52.4) | 10 (47.5) |  |  |
| **Relationship status** |  |  |  |  |
| Single | 91 (52.6) | 82 (47.4) | 1.902 | 0.079 |
| Non-Single | 14 (36.8) | 24 (63.2) | (0.923-3.922) |  |
| **Family income (monthly** |  |  |  |  |
| ≤ 30000 | 39 (59.1) | 27 (40.9) | 1.729 | 0.068 |
| More than 30000 | 66 (45.5) | 79 (54.5) | (0.959-3.118) |  |
| **Field of study** |  |  |  |  |
| MBBS/ BDS | 40 (47.6) | 44 (52.4) | NA | 0.510 |
| BAMS | 12 (46.2) | 14 (53.8) |  |  |
| BPH | 12 (42.9) | 16 (57.1) |  |  |
| BSc. Nursing | 13 (46.4) | 15 (53.6) |  |  |
| BSc. Pharmacy | 10 (55.6) | 8 (44.4) |  |  |
| Others (BMLT, BMIT,<br>BASLP and B. Perfusion<br>Technology) | 18 (66.7) | 9 (33.3) |  |  |
| **Year of study** |  |  | NA | 0.673 |
| First year | 26 (57.8) | 19 (42.2) |  |  |
| Second year | 26 (49.1) | 27 (50.9) |  |  |
| Third year | 20 (46.5) | 23 (53.5) |  |  |
| Fourth year and above | 33 (47.1) | 37 (52.9) |  |  |
| **Duration of use (per**<br>**day)** |  |  |  |  |
| ≤ 5 hours | 46 (63.9) | 26 (36.1) | 2.399 | 0.003* |
| More than 10 hours | 59 (42.4) | 80 (57.6) | (1.334-4.314) |  |
| **Internet speed** |  |  |  |  |
| ≤ 5sec | 63 (52.9) | 56 (47.1) | 1.339 | 0.332 |
| More than 5 sec | 42 (45.7) | 50 (54.3) | (0.776-2.312) |  |
| **Purpose of Use** |  |  |  |  |
| Social networking | 99(49.5) | 101 (50.5) |  |  |
| Online gaming | 36 (49.3) | 37 (50.7) |  |  |
| Entertainment | 91 (48.4) | 97 (51.6) | NA | 0.834 |
| Educational purpose | 71 (50.0) | 71 (50.0) |  |  |
| Sexually explicit content | 5 (38.5) | 8 (61.5) |  |  |

\*Significant difference at p<0.01

## Discussion

This study was conducted to assess internet addiction among undergraduate students and identify its associated factors. Approximately 50% of the undergraduates had internet addiction, of whom 36.5% had mild, 12.3% had moderate, and 1.4% had severe internet addiction. These findings are comparable to a study conducted among a similar population in India, which reported 14.4% moderate and 1.3% severe internet addiction (14). Similarly, studies conducted across six Asian countries reported prevalence rates of 14–51% using the Internet Addiction Test (IAT) with a cutoff score of ≥70 (2), while a European study across nine countries reported rates of 14–55% for moderate to severe internet addiction (15). These findings are consistent with the present study. The prevalence of severe internet addiction (1.4%) was also similar to previous studies reporting 1.3% and 1.4%, respectively (6,14). However, the prevalence of moderate internet addiction was lower than that reported in previous studies conducted in Nepal, where approximately 40% of participants had moderate addiction (6,7). This difference may be attributed to methodological variations and differences in the study population and geographical coverage.

When the major activities carried out by students were analyzed, the internet was mainly used for social networking (32.5%), information retrieval (30.5%), online gaming (23.0%), entertainment (11.9%), and sexual content (2.1%). These findings are consistent with studies conducted among similar populations in Nepal, India, and other Asian countries, suggesting that the purposes of internet use are similar among university students internationally (2,6,11,14).

Regarding the association between internet addiction and socio-demographic variables, no significant association was observed. However, studies conducted in Greece, Bangladesh, and India have reported associations between internet addiction and socio-demographic factors such as age, gender, residence, and relationship status (1,8,14). In the present study, gender was not significantly associated with internet addiction, which is consistent with a study conducted in Bangladesh (8), but contrasts with studies among medical students in India, where gender was significantly associated with internet addiction (14,16). Similarly, a study among medical students in Guilan reported significant associations with age, gender, marital status, and level of study (3). These differences suggest that the relationship between socio-demographic factors and internet addiction may vary across populations and should be further explored. Monthly family income was also not significantly associated with internet addiction, consistent with findings from India (14), whereas a study in Bangladesh reported a significant association, possibly due to differences in the study population, which comprised job-seeking graduates (8). Duration of internet use was significantly associated with internet addiction in this study, which is consistent with studies conducted in Bangladesh and India (8,14). However, internet speed was not significantly associated with internet addiction, in contrast to previous studies conducted in Nepal and India (11,14,16). This difference may be attributed to the subjective assessment of internet speed in both studies.

Internet addiction has been associated with various excessive online behaviors, including academic use, social networking, online gaming, accessing movies and music, and viewing sexually explicit content (8). Although previous studies have reported associations between internet-use activities and internet addiction (1,6), this study did not find any significant association between the purpose of internet use and internet addiction.

The bivariate analysis of academic characteristics, including year of study and field of study, showed no significant association with internet addiction. However, previous studies have reported that level of study and field of study may influence internet addiction (3,5). This difference may be due to variations in curriculum and academic activities across different fields and levels of study. The lack of significant association in this study may also be attributed to the relatively small sample size.

The study had some limitations, including the inability to include a larger number of students due to time constraints and unfavorable circumstances. In addition, the findings relied on self-reported data, which may be influenced by participants’ honesty and accuracy. As internet addiction was assessed using the self-rating Young’s Internet Addiction Test (YIAT) rather than a professional clinical diagnosis or face-to-face assessment, there is a possibility of underestimation or overestimation of internet addiction.

## Conclusion

This study was conducted to assess internet addiction among undergraduate students of IOM and identify its associated factors. Approximately half of the students had internet addiction, with more than one-tenth experiencing moderate to severe addiction. Duration of internet use per day was the only factor significantly associated with internet addiction. Therefore, rational use of the internet should be encouraged among undergraduate students. No significant association was found between internet addiction and socio-demographic or academic variables. The findings of this study may be useful for future studies to gain further insight into internet addiction among young people in Nepal.

## Abbreviations

BAMS: Bachelor in Ayurvedic Medicine and Surgery
BASLP: Bachelor in Audiology & Speech Language Pathology
BMLT: Bachelor in Medical Laboratory Technology
BMIT: Bachelor in Medical Imaging Technology
B.Pharma: Bachelor in Pharmacy
BPH: Bachelor in Public Health
BSc Nursing: Bachelor of Science in Nursing
B.Optom: Bachelor in Optometry
BDS: Bachelor in Dental Surgery
CDPH: Central Department of Public Health
CI: Confidence Interval
DSM: Diagnostic and Statistical Manual of Mental Disorders
IA: Internet Addiction
IOM: Institute of Medicine
IRC: Institutional Review Committee
MBBS: Bachelor in Medicine and Surgery
SPSS: Statistical Package for the Social Sciences
TU: Tribhuvan University
YIAT: Young’s Internet Addiction Test.

## Data availability

The data are available from the corresponding author upon reasonable request.

## Acknowledgement

We sincerely thank all the students from different faculties who participated in the study and acknowledge the Central Department of Public Health, Institute of Medicine, Nepal, for its institutional support.

## Author information

### Contributions

RL conceived the concept and design of the study. RL also conducted the survey and, with the support of SMS, conducted data analysis. SMS supervised the whole study process. All the authors have read, reviewed, and endorsed the final version of the manuscript.

## Ethics declarations

### Ethical approval and consent to participate

The study protocol was reviewed and approved by the Institutional Review Committee (IRC) of the Institute of Medicine (ref. no.: 248(6-11) E2 077/078). The research was conducted in accordance with the ethical standards of the institutional and/or national research committee and with the 1964 Helsinki Declaration and its later amendments or comparable ethical standards. Informed consent was obtained from all individual participants included in the study.

### Consent for publication

Not applicable.

